# Outpatient treatment of Covid-19 with metformin, ivermectin, and fluvoxamine and the development of Long Covid over 10-month follow-up

**DOI:** 10.1101/2022.12.21.22283753

**Authors:** Carolyn T. Bramante, John B. Buse, David Liebovitz, Jacinda Nicklas, Michael A. Puskarich, Ken Cohen, Hrishikesh Belani, Blake Anderson, Jared D. Huling, Christopher Tignanelli, Jennifer Thompson, Matthew Pullen, Lianne Siegel, Jennifer Proper, David J. Odde, Nichole Klatt, Nancy Sherwood, Sarah Lindberg, Esteban Lemus Wirtz, Amy Karger, Kenny Beckman, Spencer Erickson, Sarah Fenno, Katrina Hartman, Michael Rose, Barkha Patel, Gwendolyn Griffiths, Neeta Bhat, Thomas A. Murray, David R. Boulware

## Abstract

**Background:** Long Covid is an emerging chronic illness potentially affecting millions, sometimes preventing the ability to work or participate in normal daily activities. COVID-OUT was an investigator-initiated, multi-site, phase 3, randomized, quadruple-blinded placebo-controlled clinical trial (NCT04510194). The design simultaneously assessed three oral medications (metformin, ivermectin, fluvoxamine) using two by three parallel treatment factorial assignment to efficiently share placebo controls and assessed Long Covid outcomes for 10 months to understand whether early outpatient treatment of SARS-CoV-2 with metformin, ivermectin, or fluvoxamine prevents Long Covid.

**Methods:** This was a decentralized, remotely delivered trial in the US of 1,125 adults age 30 to 85 with overweight or obesity, fewer than 7 days of symptoms, and enrolled within three days of a documented SARS-CoV-2 infection. Immediate release metformin titrated over 6 days to 1,500mg per day 14 days total; ivermectin 430mcg/kg/day for 3 days; fluvoxamine, 50mg on day one then 50mg twice daily through 14 days. Medical-provider diagnosis of Long Covid, reported by participant by day 300 after randomization was a pre-specified secondary outcome; the primary outcome of the trial was severe Covid by day 14.

**Result:** The median age was 45 years (IQR 37 to 54), 56% female of whom 7% were pregnant. Two percent identified as Native American; 3.7% as Asian; 7.4% as Black/African American; 82.8% as white; and 12.7% as Hispanic/Latino. The median BMI was 29.8 kg/m^2^ (IQR 27 to 34); 51% had a BMI >30kg/m^2^. Overall, 8.4% reported having received a diagnosis of Long Covid from a medical provider: 6.3% in the metformin group and 10.6% in the metformin control; 8.0% in the ivermectin group and 8.1% in the ivermectin control; and 10.1% in the fluvoxamine group and 7.5% in the fluvoxamine control. The Hazard Ratio (HR) for Long Covid in the metformin group versus control was 0.58 (95% CI 0.38 to 0.88); 0.99 (95% CI 0.592 to 1.643) in the ivermectin group; and 1.36 in the fluvoxamine group (95% CI 0.785 to 2.385).

**Conclusions:** There was a 42% relative decrease in the incidence of Long Covid in the metformin group compared to its blinded control in a secondary outcome of this randomized phase 3 trial.

**Trial registration:** NCT04510194.

**IND:** 152439

## Background

Infection with severe-acute respiratory coronavirus 2 (SARS-CoV-2) has been observed to cause Post-Acute Sequelae of Covid (PASC), commonly referred to as “Long Covid.”^1^ The experience of Long Covid is heterogenous, ranging from a single symptom to serious multi-organ involvement, and from mild and short lived to chronically debilitating.^1,2^ The Centers for Disease Control and Prevention (CDC) estimates that Long Covid disproportionately affects racial and ethnic minority populations, which makes understanding and reducing the incidence of Long Covid critically important.^1,3^

Cross-sectional studies estimate that 15% of adults in the US have symptoms after SARS-CoV-2 infection that correlate with a diagnosis of Long Covid.^4^ One of the largest prospective cohorts to study persistent symptoms after Covid-19 suggests that somatic symptoms could be attributable to SARS-CoV-2 in approximately 12% of adults in the cohort.^5^ An important gap in the literature is understanding the proportion of adults infected with SARS-

CoV-2 who are diagnosed with Long Covid by medical providers. Previous efforts have tried to understand Long Covid using electronic health record data, but reliably capturing the condition is challenging.^6,7^ A code in the International Classification of Diseases, 10^th^ Edition, was not added until October 2021, and there are concerns about its sensitivity and specificity.^1,8,9^

COVID-OUT was a phase 3 randomized, quadruple-blinded placebo-controlled trial of early outpatient treatment of SARS-CoV-2 that included monthly follow-up for 300 days to test the hypothesis that early treatment of Covid-19 with the study drugs would prevent Long Covid.^10^ The trial used a 2 by 3 factorial design of parallel treatments to assess: metformin, ivermectin, and/or fluvoxamine. Comparing the incidence of Long Covid between treatment and randomized control groupss was a prespecified secondary outcome.

## Methods

### Trial Design and Oversight

COVID-OUT was an investigator-initiated, multi-site, phase 3, randomized, quadruple-blinded placebo-controlled clinical trial (ClinicalTrials.gov: NCT04510194).^10^ Those blinded included: participants, care providers, investigators, and outcomes assessors. The trial was decentralized, with no in-person contact with participants. Informed consent was obtained from each participant via electronic consent, or written consent if they did not have an email address.

Institutional review boards at each site, and Advara centrally, approved the protocol. An independent data safety monitoring board oversaw safety and efficacy monitoring, and an independent monitor oversaw study conduct in compliance with the Declaration of Helsinki, Good Clinical Practice Guidelines, and local requirements.^11^

### Intervention and Randomization

The trial design simultaneously assessed three distinct oral medications (metformin, ivermectin, fluvoxamine) using a two by three parallel treatment factorial assignment to efficiently share placebo controls. Participants were randomized 1:1:1:1:1:1 to each study arm as described in a previous publication and shown in **Figure 1**.^10^ Briefly, the trial opened with a 1:1 randomization to metformin versus placebo on December 30, 2020 and the factorial design opened May 21, 2021. Enrollment ended January 28, 2022 and all investigators except the unblinded statistician remained blinded to group-level results through February 14, 2022. The Day 300 follow-up ended Nov 27, 2022. All investigators, outcome assessors, treating clinicians, and participants remain blinded to individual treatment allocations.

**Figure 1.**
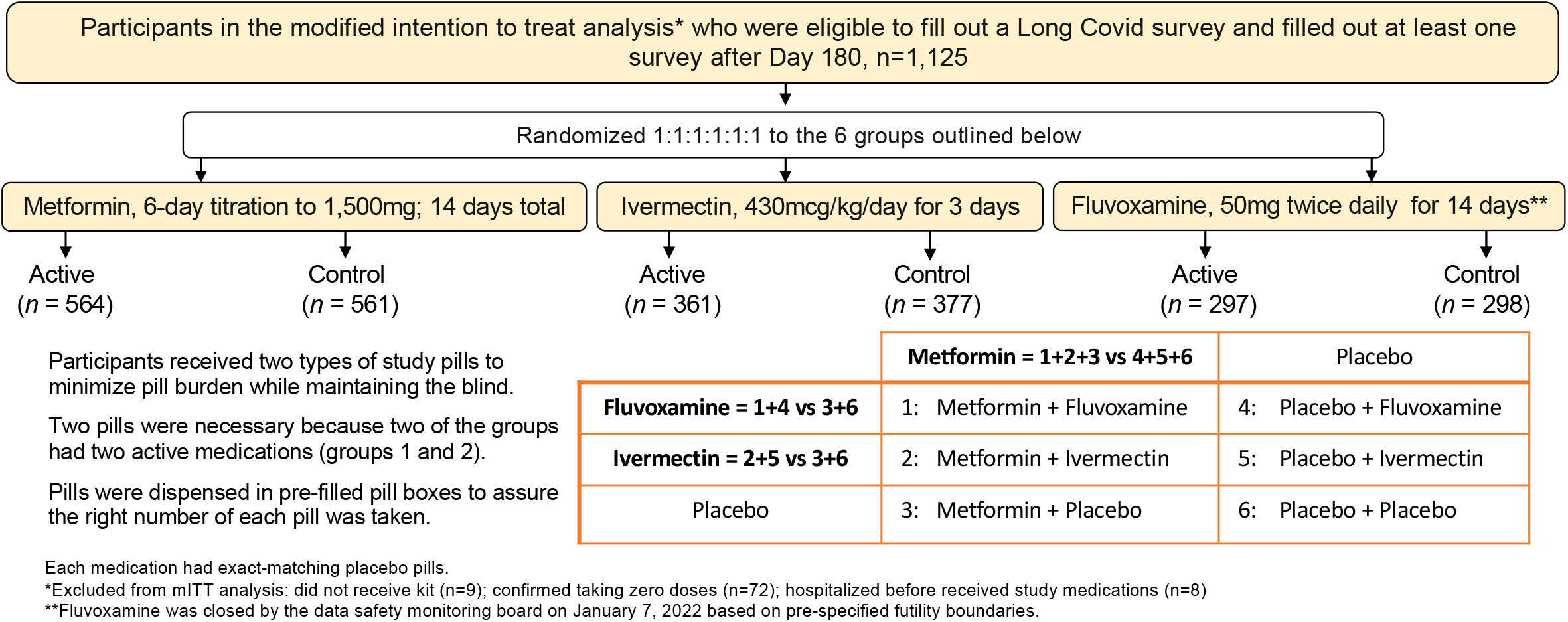
Overview of factorial design groups. Participants were randomized to equal allocation to one of the 6 arms above. They were compared to concurrently randomized controls from the groups outlined above. Every participant in the trial received a pill that looked like metformin – either active metformin or exact matching metformin placebo.

**Figure 2.**
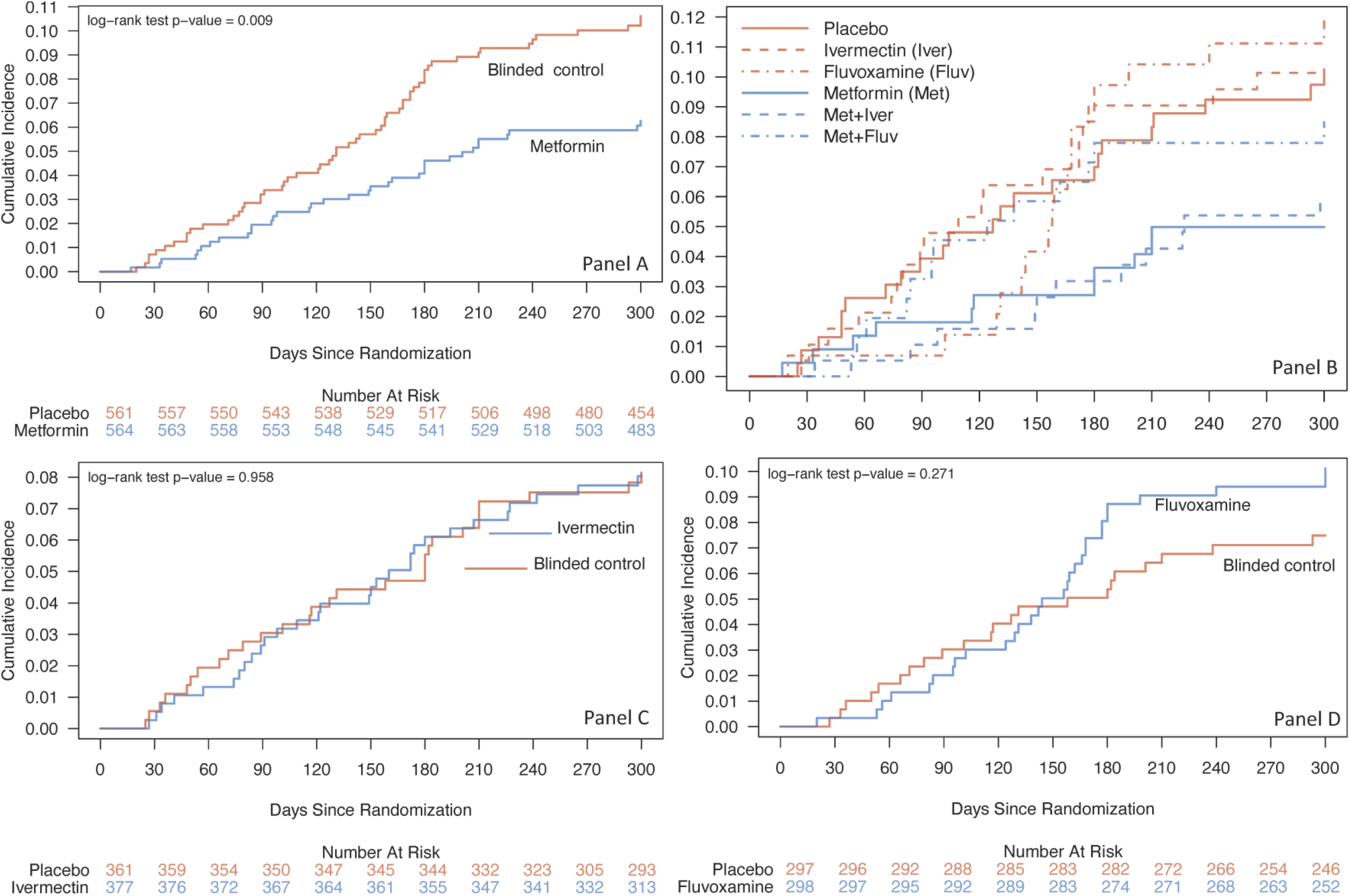
Cumulative incidence of “Long Covid,” post-acute sequelae of SARS-CoV-2 infection (PASC), diagnosed by a medical provider over 10 months after randomization. Panel A is metformin; Panel B is all 6 arms in the trial; Panel C is ivermectin; Panel D is fluvoxamine. In this factorial design trial, each participant received two types of pills. Every participant received a pill that looked like metformin -- either metformin or an exact-matching metformin placebo. The second pill was active fluvoxamine or ivermectin, or their exact matching placebo.

Manufacturers provided exact-matching placebo pills and in the factorial design each participant received two types of pills to maintain the blind: 1) metformin or exact-matching metformin placebo; and 2) fluvoxamine, ivermectin, or their exact-matching placebo. The medications were pre-packaged into pill boxes to speed delivery to participants and assure participants took the correct number of each type of pill. Study medication was sent via same-day courier or overnight shipping to participants. The average time from consent to receipt of study drug was <1 day. The metformin dose was titrated over 6 days: 500mg on day 1; 500mg twice daily for 4 days; then 500mg mornings and 1000mg evening through 14 days. The ivermectin dose was 390-470 mcg/kg per day for 3 days (median 430 mcg/kg/day). Fluvoxamine was 50mg on Day 1 followed by 50mg twice daily through 14 days.

### Outcomes

To understand whether metformin, ivermectin, or fluvoxamine prevent the development of Long Covid, participants were asked whether they had any new diagnoses since their infection with SARS-CoV-2. Specific questions included whether a medical provider gave them a diagnosis of Long Covid, and if so when and what type of provider gave this diagnosis.

Because the definition of Long Covid is rapidly changing, fluctuating symptoms are challenging to assess, and ICD codes lack specificity and sensitivity, the primary method for ascertaining Long Covid was participant-reported receipt of a Long Covid diagnosis from a medical provider. Medical providers and patients are wary of using diagnosis labels in the medical record unnecessarily, because doing so can negatively impact patient psyche and have insurance ramifications. Participants consented to medical record review for confirmation of these diagnoses.

Participants were also asked about SARS-CoV-2 re-infection, severity of reinfection, and vaccination against SARS-CoV-2 after randomization. Survey questions are available in the **Supplement**.

### Participants

Eligibility criteria included: age 30 to 85 years with overweight or obesity by self-reported body mass index (BMI); documentation of confirmed SARS-CoV-2 infection; <7 days of symptoms; and no known prior infection with SARS-CoV-2. Participants had to provide consent within 3 days of their positive SARS-CoV-2 test. Participants were excluded if they were already taking one of the study medications or if they had already received an EUA-approved Covid-19 treatment. Home medications and treatments received after enrollment were recorded. Vaccination against SARS-CoV-2 was not an exclusion criterion.

Pregnant and lactating women were included and were randomized 1:1 to metformin or placebo, not fluvoxamine or ivermectin due to less established literature for safety during pregnancy and lactation for those medications.

### Trial Procedures

The active follow-up period for the trial was 28 days. Beginning at 60 days post randomization, surveys were sent every 30 days through 300 days (10 months) after randomization via automated email or other per patient preference. Ten-month follow-up for Long Covid was not in the original protocol as Long Covid was not a known entity in fall 2020.

The pre-specified secondary endpoint on Long Covid was added to the protocol in April 2021, and survey tools were IRB-approved in July 2021. Participants enrolled before the Long Covid surveys were approved were contacted for reconsent to receive the Day 300 Long Covid survey. An overview of protocol changes is in the **Supplement**.

### Statistical Analysis

A factorial, 2 by 3 design of distinct, parallel treatments with exact-matching placebo pills allows the simultaneous conduct of three separate randomized trials that efficiently share concurrently randomized, blinded controls. Correcting for multiple comparisons for a factorial design is not appropriate.^12,13^ Accordingly, factorial design trials often publish each medication in the trial separately.^14-17^

The comparison groups for each study drug consists of persons who were assigned the active version of the drug versus those who were at risk of being assigned to the active version of the drug but were assigned a blinded control instead (**Figure 1)**. By design, the active and control comparison groups have balanced numbers of persons receiving active and placebo version of the other study drug.

The *a priori* primary sample population was a modified intention to treat (mITT) sample. Participants who did not receive the study medication; were hospitalized at the time of delivery; or reported not taking any study doses were excluded from the mITT.^10^ The Long Covid questionnaire and consent was added after July, 2021, so within the mITT sample (n=1,323) this analysis was restricted to the 1,125 participants who consented to long term follow up and completed at least 1 survey on or after Day 180 (**Figure 1)**.

Reports of Long Covid diagnosis by medical provider were analyzed using a time-to-event approach with time denoting the time from randomization. This approach appropriately accounts for participants who did not fill out all the potential Long Covid surveys, and thus were lost to follow up prior to Day 300. For persons who reported a Long Covid diagnosis, the date of their diagnosis was set to the 15th day of the earliest month in which they reported receiving the diagnosis. For persons who reported a Long Covid diagnosis but did not provide valid timing of diagnosis information (n=9), (i.e. they provided a month where the last day in that month occurred earlier than 15 days from their randomization) the date of their diagnosis was set to the study day of the earliest Long Covid survey on which they reported the diagnosis. Participants who did not report a Long Covid diagnosis were censored based on the study day of their latest Long Covid survey. A time-to-event approach also adds knowledge about this new disease state by reporting when individuals are receiving diagnoses of Long Covid.

## Results

### Study Participants

Overall 1,125 consented for Long Covid follow-up and completed at least one survey on or after Day 180, 564 in the metformin group and 561 in the blinded control group. The median age was 45 years (IQR 37 to 54), 56% were female of whom 7% were pregnant. Two percent identified as Native American; 3.7% as Asian; 7.4% as Black/African American; 82.8% as white; and 12.7% as Hispanic/Latino. The median BMI was 29.8 kg/m^2^ (IQR 27.0 to 34.2), and 51% had a BMI >30kg/m^2^. The median days from symptom onset to study drug initiation was 5 days (IQR 4 to 6), and 46.8% started study drug within 4 days or less of symptom onset. Overall, 55% (n=618) had received the primary Covid-19 vaccination series, including 5.1% (n=57) who received a booster, before enrollment (**Table 1)**.

**Table 1:**
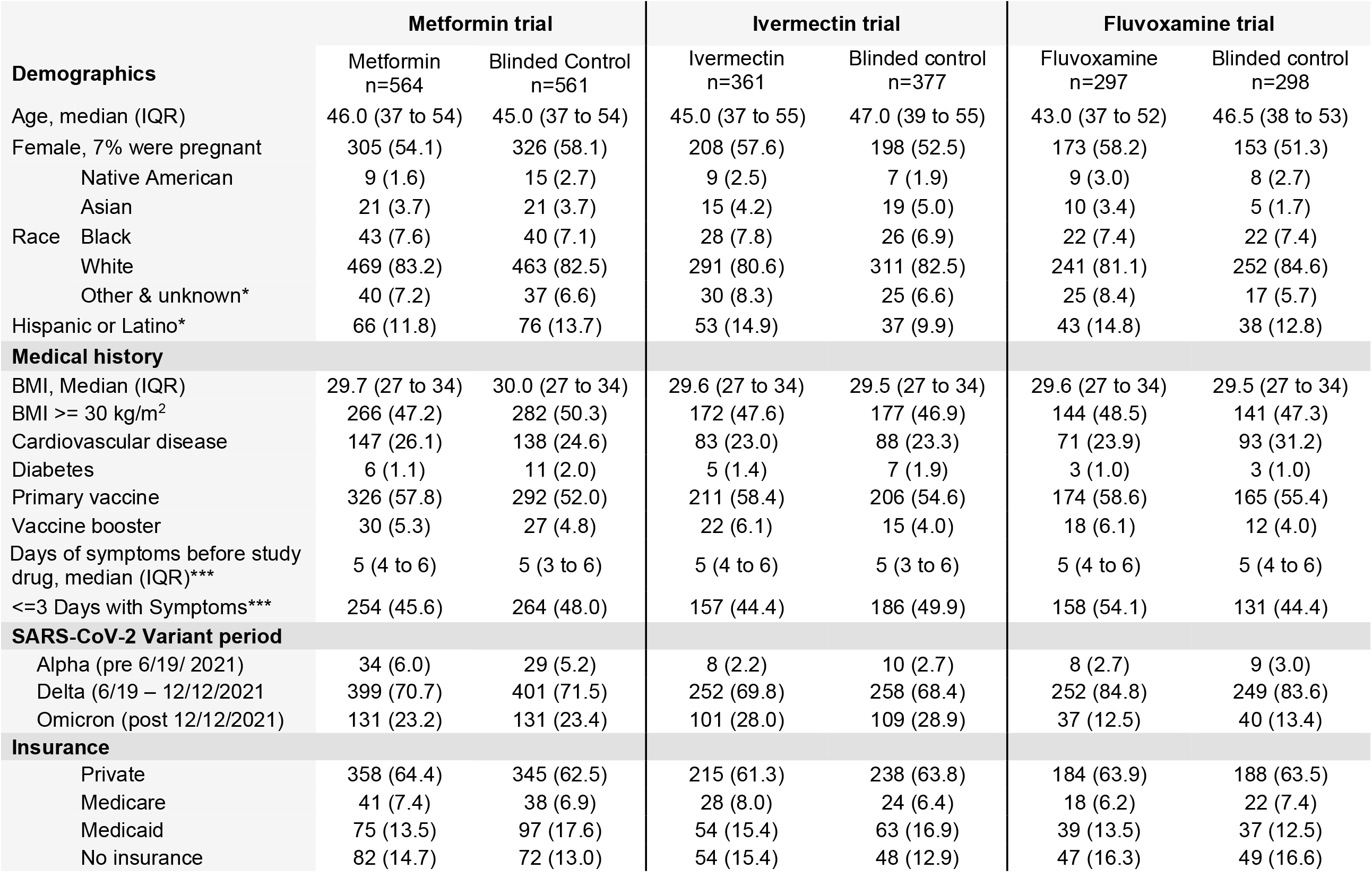
Baseline characteristics by randomized treatment groups. Values are n (%) unless specified. Abbreviations: BMI = body mass index; IQR=inter-quartile range; SD = standard deviation. Cardiovascular disease defined as: hypertension, hyperlipidemia, coronary artery disease, past myocardial infarction, congestive heart failure, pacemaker, arrhythmias, or pulmonary hypertension. *Includes Hawaiian / Pacific Islander. **missing n=9 with Hispanic ethnicity information. ***missing symptom duration for n=18.

### Long Covid Diagnosis

Overall, 8.4% (94/1125) responded Yes to the question: “Has a medical provider told you that you have Long Covid?” Most of the Long Covid diagnoses were made by primary care providers, n=72 (73.4%); followed by a provider specializing in Long Covid, n=4 (4.3%); other specialists, n=8 (cardiology n=3, neurology n=1, infectious disease n=1, otolaryngologist n=1, pulmonologist n=1); emergency department n=3; in a hospital n=2; urgent care n=2; 1 by chiropractor; 1 other; 1 missing.

Among those randomized to metformin the cumulative incidence for developing Long Covid was 6.2% (95% CI 4.2% to 8.2%), and 10.6% (8.0% to 13.1) in the blinded controls (**Figure 1, Table 2)**. For those randomized to ivermectin, the cumulative incidence was 8.0% (95% CI 5.2% to 10.8%) and 7.5% (95% CI 4.7% to 10.2%) in blinded controls (**Supplemental Table 2**). Among those randomized to fluvoxamine, the cumulative incidence was 10.1% (95% CI 6.6% to 13.5%), and 7.5% (95% CI 4.4% to 10.5% in the blinded controls (**Supplemental Table 2**).

**Table 2.** Cumulative incidence of Long Covid, percent with 95% confidence intervals.

| Day | Metformin | Blinded Control | Absolute Risk Reduction | Percent completing surveys |  |
| --- | --- | --- | --- | --- | --- |
|  |  |  |  | Metformin | Blinded control |
| **60** | 1.1%<br>(0.2% - 1.9%) | 2.0%<br>(0.8% - 3.1%) | 0.9%<br>(2.3% to -0.5%) | 99% | 98% |
| **120** | 2.8%<br>(1.5% - 4.2%) | 4.1%<br>(2.4% - 5.7%) | 1.3%<br>(3.4% to -0.9%) | 97% | 96% |
| **180** | 4.6%<br>(2.9% - 6.3%) | 8.4%<br>(6.1% - 10.6%) | 3.8%<br>(6.6% to 0.9%) | 96% | 92% |
| **240** | 5.9%<br>(3.9% - 7.8%) | 9.6%<br>(7.2% - 12.1%) | 3.8%<br>(6.9% to 0.7%) | 92% | 89% |
| **300** | 6.3%<br>(4.2% - 8.2%) | 10.6%<br>(8.0% - 13.1%) | 4.4%<br>(7.6% to 1.1%) | 86% | 81% |
| **Hazard Ratio = 0.576 (95% CI 0.379 to 0.875)** |  |  |  |  |  |

In this 2x3 factorial design randomized trial, each participant received two types of study pills: 1) a study pill that looked like metformin -- either metformin or an exact-matching metformin placebo; and 2) active flvoxamine or ivermectin, or their exact matching placebo. The hazard ratio after adjusting for the other study drugs and vaccination was 0.588 (95% CI 0.387 to 0.894).

For the metformin versus blinded control, the Hazard Ratio (HR) for developing Long Covid was 0.58 (95% CI 0.38 to 0.88); for ivermectin was HR 0.99 (0.59 to 1.64); and for fluvoxamine was HR 1.36 (0.79 to 2.39) (**Supplemental Table 2)**. Heterogeneity of treatment effect was assessed for metformin across a priori subgroups of baseline risk factors (**Figure 3)**. The effect of metformin for preventing Long Covid was consistent across subgroups, including across other study drugs and viral variants.

**Figure 3.**
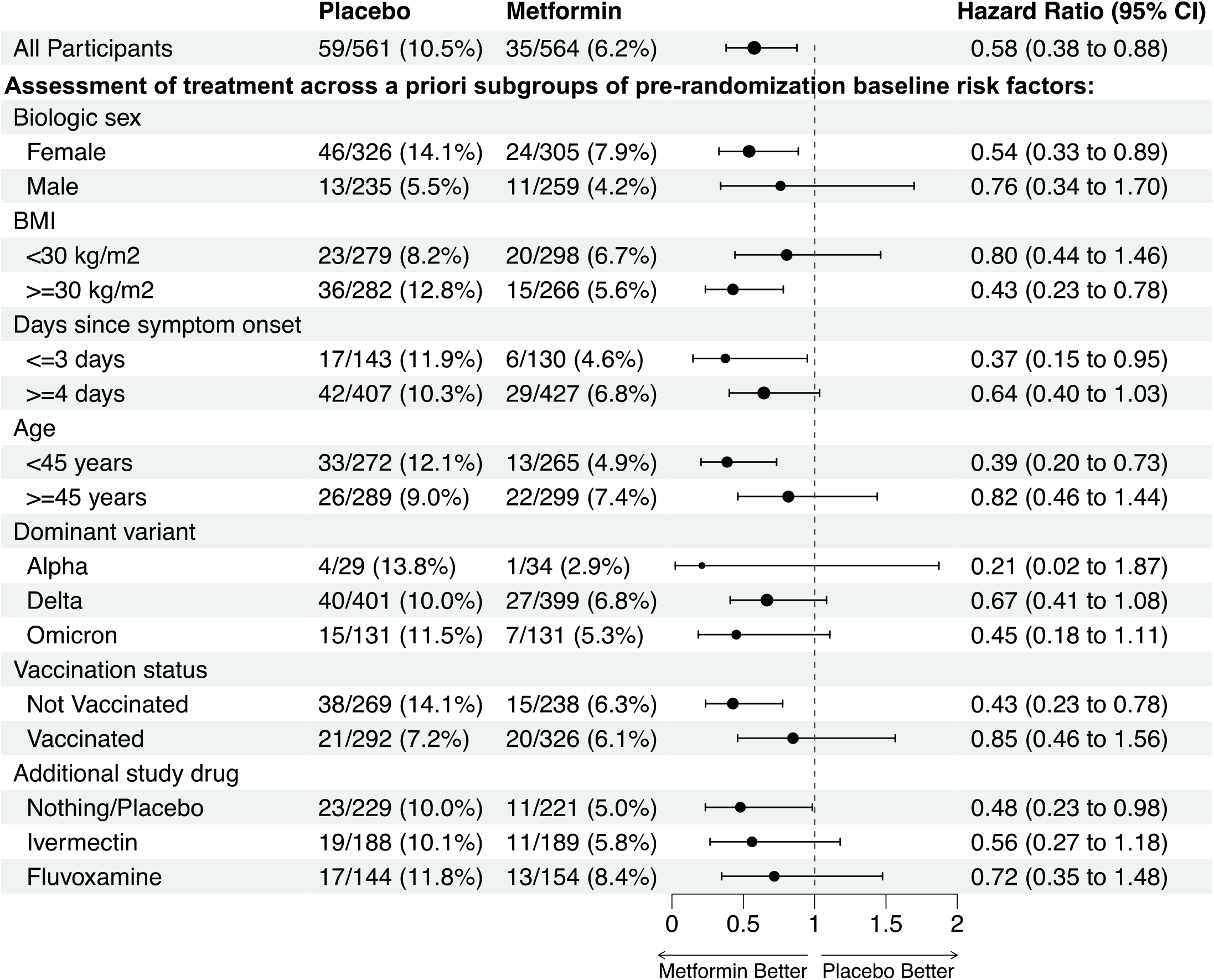
Assessment of heterogeneity of Metformin treatment effect for preventing the development of Long Covid compared to blinded control. In this factorial design trial, each participant received two types of pills. Every participant received a pill that looked like metformin -- either metformin or an exact-matching metformin placebo. The second pill was active fluvoxamine or ivermectin, or exact matching placebo for fluvoxamine or ivermectin. Fluvoxamine enrollment halted on January 7, 2022 by the independent data and safety monitoring board. Overall trial enrollment completed on January 28, 2022 when reaching the target sample size.

**Supplemental Table 2** shows proportion of participants who developed Long Covid by baseline risk factors.

## Discussion

COVID-OUT was an investigator-initiated, multi-site, phase 3, randomized, quadruple-blinded, placebo-controlled clinical trial that assessed the incidence of Long Covid diagnoses made by a healthcare provider over the 300 days following randomization. Overall, 10.6% of blinded control participants reported receiving a diagnosis of Long Covid from a medical provider, compared to 6.3% receiving metformin. The hazard ratio of 0.58 (95% CI 0.38 to 0.88) for metformin preventing Long Covid is consistent with results in the acute phase of the trial, in which metformin showed a consistent direction of effect for preventing ED visits, hospitalizations, or death due to Covid-19 (OR 0.58, 95% CI 0.35 to 0.94), and hospitalizations or death (OR 0.47, 95% CI 0.20 to 1.11) through Day 14.^10,18^ There was no decreased incidence of Long Covid attributable to ivermectin or fluvoxamine in this trial.

Factorial design trials assessing parallel distinct treatments are often misunderstood as needing correction for multiplicity for the multiple treatments. However, such design is an efficient way to simultaneously test separate interventions while sharing a control group. Platform trials also assess multiple treatments, but not necessarily simultaneously. Correcting for multiple comparisons for a factorial design is not necessary or appropriate.^12,13^ Factorial design trials often publish each medication in the trial separately, so that each medication has the depth of analysis that is expected from phase 3 randomized trials: secondary outcomes, heterogeneity of treatment effect, and even post-hoc analyses all in the main outcomes paper.^14-17^ It is important to consider all such analyses when critically evaluating a phase 3 randomized trial.^18,19^

Long Covid was a pre-specified secondary outcome of the COVID-OUT trial. It was not possible to elevate it to co-primary outcome because the trial had already begun enrolling before Long Covid was recognized as a disease entity that needed further study, and primary outcomes cannot be changed after enrollment starts. Like other trials during the Covid-19 pandemic, it was important to publish short term results before long-term follow-up was complete,^20^ and to present hypothesis testing of prespecified secondary outcomes.^20^ The p value (0.009) for metformin preventing Long Covid is low enough that it would still be less than 0.05 after applying a Bonferonni correction for the multiple testing of the primary and all four secondary clinical outcomes in this trial.

The exact pathophysiology of Long Covid is unknown but is likely multi-factorial, including the inflammatory cascade during acute infection and persistent viral replication.^21^ Mechanistic in silico modeling predicts that translation of SARS-CoV-2 viral proteins is an especially sensitive target for inhibition, and previous studies show metformin capable of suppressing protein translation via mTor.^22,23^ Experimentally, metformin has in-vitro activity at a physiologically relevant dose against SARS-CoV-2 in cell culture and in human lung tissue, ex vivo, and in a phase 2 clinical trial.^24-27^

In addition to in vitro and in vivo activity against SARS-CoV-2, metformin has been extensively studied for its anti-inflammatory actions. In human bronchial and lung epithelial cell lines infected with SARS-CoV-2, metformin restored autophagic flux, inhibited cleavage of caspase-1 by non-structural protein 6 (NSP6), and inhibited maturation and release of interleukin-1*β* and interleukin-18.^28^ Metformin also prevented a senescent phenotype induced by SARS-CoV-2 infection in dopaminergic neurons in vitro, which could be relevant to neurocognitive sequelae of infection seen in Long Covid.^29^

Several factors could influence whether an individual receives a diagnosis of Long Covid from a medical provider, such as access to medical care, competing demands that prevent receiving medical care, and willingness to seek medical care for post-Covid symptoms. These factors may introduce selection bias around who is receiving a diagnosis of Long Covid. Such factors would be expected to be equally distributed between treatment arms by the randomization in this clinical trial and should not influence our interpretation of results. In **Supplement Table 1** we present baseline characteristics comparing those who reported a diagnosis of Long Covid and those who did not.

Metformin is a safe medication. Safety concerns have centered around a risk of lactic acidosis, but that historical concern was largely driven by experience with other biguanides. Several large studies and Cochrane reviews have demonstrated no increased risk of lactic acidosis, and in fact fewer cases of lactic acidosis, in persons on metformin.^30,31^ Metformin is also safe in adults with kidney disease and should not be withheld from persons with glomerular filtration rates >30ml/min/1.73m^2^, and perhaps even lower, because of associations with improved macrovascular outcomes in persons with chronic kidney disease.^30^ Guidelines recommend metformin should no longer be stopped upon hospital admission or for surgery.^32-35^ Metformin’s safety has been demonstrated in children and during pregnancy.^36-41^

Future research is needed to understand optimal dosing regimens for preventing Long Covid, whether extended release is effective in persons who have side effects from immediate release metformin, and whether metformin could be used as a treatment for Long Covid. Future research could also assess whether metformin is effective if started during an emergency department visit or hospitalization for Covid-19.

### Limitations

The changing nature of the definition of Long Covid is a limitation. The use of a Long Covid diagnosis assigned by a medical provider, as well as the long duration of follow-up, would address some of the issues around the changing nature of this disease definition. The quadruple blinded, randomized nature of the trial limits potential biases compared to observational cohorts or case-control studies. Other non-specific events not caused by COVID-19 would be equally distributed by randomized trial arm. This trial excluded low-risk individuals: those with a normal BMI and age < 30 years, and whether these findings would generalize to those populations is unknown. Additionally, it is unknown if these findings would generalize to early outpatient treatment of SARS-CoV-2 in someone who had previously been infected with SARS-CoV-2. The sample of participants in this trial was mostly white (82.8%), compared to 76% of the US population; and only 12.7% identified as Latino or Hispanic.^42^ With 56% of participants being female, it was well balanced by sex. While only 7% of females in the trial were pregnant, this was one of few randomized trials of outpatient Covid-19 treatment to enroll pregnant women.^10,43^

## Conclusions

Among adults with Covid-19, outpatient treatment with metformin decreased the development of Long Covid by 42% in a phase 3 randomized trial whose sample was mostly vaccinated and included enrollment during the Omicron wave. This is notable because Long Covid is a significant public health emergency that may have lasting health, mental health, and economic sequelae, especially in socioeconomically marginalized groups. This finding is consistent with the observed 42% reduction in emergency department visits, hospitalizations, or death with metformin in the first 14 days of the trial; and reduced hospitalization or death in the first 28 days, during which 1.34% (8/596) of those receiving metformin were hospitalized or died compared to 3.16% (19/601) of blinded controls. Further clinical trials are warranted to assess whether there is synergy with other treatments, such as nirmatrelvir in vaccinated populations or in those with prior Covid-19. Fluvoxamine and ivermectin did not decrease the development of Long Covid, which is consistent with outcomes in the first 14 days of the trial.

## Data Availability

All data produced in the present study are available within 2 weeks of request.
All data used to produce the 2-week outcomes paper is already available on covidout.umn.edu

https://covidout.umn.edu

## Acknowledgements

We thank the participants in the trial. We would also like to thank many others who made this trial possible, including: The M Health Fairview Obesity Medicine Research Advisory Panel, particularly Stacy Dean and Yelena Kibasova. The numerous volunteers who helped fold and tape boxes and place labels so that the study team could focus on enrollment and follow-up.

Volunteers include: Stacy Washington, Ben Tsech, Sasha Fraser, Evan Fraser, Piotr Bednarski, Paloma Good, Josie June Veit.

University of Minnesota, M Health Fairview: Program in Health Disparities Research; Clinical and Translational Science Institute’s (CTSI) Best Practices Integrated Informatics Core (BPIC); Medical School Communications; M Health Fairview Recruiting Office; Department of Surgery Clinical Trials Office; Fairview Investigational Drug Services Pharmacy; Sponsored Projects Administration; Advanced Research and Diagnostic Laboratory; Center for Pediatric Obesity Medicine; UMN Institute for Engineering in Medicine; CTSI Regulatory support; Department of Medicine Research Operations and Division of General internal Medicine, especially Jill Charles, Manuria Yang, and Kate Brekke.

Dr. Bramante thanks her KL2 and K23 mentors for their continued career mentorship and support: Anne Joseph, MD, MPH; Aaron Kelly, PhD; Claudia Fox, MD, MPH; and Kimberly Gudzune, MD, MPH.

Dr. Bramante thanks the M Health Fairview Learning Health Systems career development program and mentors Genevieve Melton-Meaux, MD, PhD and Bradley Benson, MD; and Fairview Research Services, especially Andrew Snyder and Jill Cordes. Dr. Bramante also thanks other colleagues and mentors who contributed to considerations for the protocol: Eric Lenze, MD; Angela Reiersen, MD; David Haynes, PhD; Carlos Chaccour, MD; Ildilko Linvay, MD; Ana Palacio, MD; Leonardo Tamariz, MD, MPH; Ananth Shalev, MD; Erik Anderson, MD; and Jeanne M. Clark, MD, MPH.

