## Supplemental material for "Outpatient treatment of Covid-19 with metformin, ivermectin, and fluvoxamine and the development of Long Covid over 10-month follow-up"

**Supplemental Table 1:** Baseline characteristics between those who reported receiving a diagnosis of Long Covid and those who reported no diagnosis of Long Covid.

| | | Overall n=1,125 | No Long Covid n=1031 (91.6%) | Long Covid n=94 (8.4%) |
|---|---|---|---|---|
| **Age, median (IQR)** | | 45.0 (37.0 to 54.0) | 45.0 (37.0 to 55.0) | 45.0 (38.0 to 51.0) |
| **Female** | | 631 (56.1) | 561 (54.4) | 70 (74.5) |
| **Pregnant** | | 44 (3.9) | 41 (4.0) | 3 (3.2) |
| **Race** | Native American | 24 (2.1) | 21 (2.0) | 3 (3.2) |
| | Asian | 42 (3.7) | 39 (3.8) | 3 (3.2) |
| | Hawaiian / Pacific Islander | 7 (0.6) | 6 (0.6) | 1 (1.1) |
| | Black | 83 (7.4) | 72 (7.0) | 11 (11.7) |
| | White | 932 (82.8) | 855 (82.9) | 77 (81.9) |
| | Other and unknown | 70 (6.2) | 68 (6.6) | 2 (2.1) |
| **Hispanic or Latino *** | | 142 (12.7) | 133 (13.0) | 9 (9.7) |
| **Medical history** | | | | |
| BMI, median (IQR) | | 29.8 (27.0 to 34.2) | 29.7 (26.8 to 33.9) | 31.0 (27.5 to 36.0) |
| BMI >= 30 kg/m$^2$ | | 548 (48.7) | 497 (48.2) | 51 (54.3) |
| Cardiovascular Disease | | 285 (25.3) | 263 (25.5) | 22 (23.4) |
| Diabetes | | 17 (1.5) | 17 (1.6) | 0 (0.0) |
| Primary vaccine before enrollment | | 618 (54.9) | 577 (56.0) | 41 (43.6) |
| Vaccine booster before enrollment | | 57 (5.1) | 56 (5.4) | 1 (1.1) |
| Any Vaccine after enrollment | | 160 (14.2) | 144 (14.0) | 16 (17.0) |
| Days of symptoms before study drug initiation, median (IQR)* | | 5 (4 to 6) | 5 (4 to 6) | 5 (4 to 6) |
| <=3 Days with Symptoms* | | 518 (46.8) | 480 (47.4) | 38 (40.4) |
| **Variant period** | | | | |
| | Alpha (before 6/19/ 2021) | 63 (5.6) | 58 (5.6) | 5 (5.3) |
| | Delta (6/19 – 12/12/2021) | 800 (71.1) | 733 (71.1) | 67 (71.3) |
| | Omicron (after 12/12/2021) | 262 (23.3) | 240 (23.3) | 22 (23.4) |
| **Insurance status** | | | | |
| | Private | 703 (63.4) | 651 (64.1) | 52 (55.9) |
| | Medicare | 79 (7.1) | 70 (6.9) | 9 (9.7) |
| | Medicaid | 172 (15.5) | 152 (15.0) | 20 (21.5) |
| | No insurance | 154 (13.9) | 142 (14.0) | 12 (12.9) |
| **Randomized to metformin** | | 564 (50.1) | 529 (51.3) | 35 (37.2) |
| **Randomized to ivermectin** | | 377 (33.5) | 347 (33.7) | 30 (31.9) |
| **Randomized to fluvoxamine** | | 298 (26.5) | 268 (26.0) | 30 (31.9) |

Values are n (%), median (interquartile range), or mean (±Standard Deviation).
Abbreviations: BMI = body mass index; IQR=inter-quartile range;
Cardiovascular disease defined as: hypertension, hyperlipidemia, coronary artery disease, past myocardial infarction, congestive heart failure, pacemaker, arrhythmias, or pulmonary hypertension.

*missing n=18 for symptom duration; missing n=9 of Hispanic ethnicity

**Supplemental Table 2. Cumulative incidence of Long Covid diagnoses.**

| Day | Blinded Control 29/361 (8.0%) | Ivermectin 30/377 (8.0%) | Ivermectin Absolute Risk Reduction | Blinded Control 22/297 (7.4%) | Fluvoxamine 30/298 (10.1%) | Fluvoxamine Absolute Risk Reduction |
|---|---|---|---|---|---|---|
| 60 | 1.9% (0.5% to 3.4%) | 1.3% (0.2% to 2.5%) | 0.6% (2.4% to -1.2%) | 1.7% (0.2% to 3.1%) | 1.0% (0.0% to 2.1%) | 0.7% (2.5% to -1.2%) |
| 120 | 3.9% (1.9% to 5.8%) | 3.4% (1.6% to 5.3%) | 0.4% (3.1% to -2.3%) | 4.0% (1.8% to 6.3%) | 3.0% (1.1% to 4.9%) | 1.0% (4.0% to -1.9%) |
| 180 | 5.5% (3.2% to 7.9%) | 6.1% (3.7% to 8.5%) | -0.6% (2.8% to -3.9%) | 5.4% (2.8% to 7.9%) | 8.7% (5.5% to 11.9%) | -3.3% (0.8% to -7.4%) |
| 240 | 7.5% (4.7% to 10.2%) | 7.2% (4.5% to 9.8%) | 0.3% (4.1% to -3.4%) | 7.1% (4.1% to 10.0%) | 9.4% (6.0% to 12.7%) | -2.3% (2.1% to -6.7%) |
| 300 | 7.5% (4.7% to 10.2%) | 8.0% (5.2% to 10.8%) | 0.1% (4.1% to -3.8%) | 7.5% (4.4% to 10.5%) | 10.1% (6.6% to 13.5%) | -2.6% (1.9% to -7.2%) |
| **Hazard Ratio = 0.986 (0.592 to 1.643)** | | | | **Hazard Ratio= 1.360 (0.785 to 2.358)** | | |

Participants were randomized to ivermectin, fluvoxamine, or identical matched placebo. Ivermectin was dosed at 390 to 470 μg per kilogram per day for 3 days (median 430 μg/kg/day), and fluvoxamine dosed at 50 mg twice daily for 14 days.

**Supplemental Table 3.**

**Outcome Ascertainment**

*Below is the question asked in monthly surveys for 9 months after completion of the acute phase of the trial (9 months after Day 28 of the trial). The questions, answer options with branching logic, that were analyzed for this manuscript are presented.*

**We have a few questions about your health since you enrolled in the study:**

**1)** Has a medical provider told you that you have "Long Covid" Yes/No

– If yes: "Approximately when? _____(month)"

– If yes: "Who Told you?"

- My primary care provider;
- A provider who specializes in Long Covid;
- A specialist; then branching logic for: cardiologist; neurologist; pulmonologist; other:_______
- A chiropractor;
- Other:_______

**Supplemental Table 4.**

**Overview of changes to the Protocol for adding assessments of Long Covid**

**Overview of changes to the Protocol for adding assessments of Long Covid with links to clinicaltrials.gov**

- The protocol version dates on the front page of each protocol.
- Only the first and final protocols were published with the first outcomes paper.
- We submit links to each version of the protocol after Long Covid was added:

  - **April, 2021,** Version 3.1: Long Covid / PASC was added as an outcome (section 3.1), initially under primary outcomes.
  - **July, 2021,** Version 3.2: Long Covid / PASC questionnaire was added as a protocol addendum
  - **Sept, 2021,** Version 3.3**:** small protocol changes, not related to PASC
  - **Dec 8, 2021**, Version 3.4: moved PASC down to secondary outcomes. This final version of the protocol was published on clinicaltrials.gov in January 20, 2022 while enrollment was still ongoing.
    - **Text in protocol version 3.4:**
      - "Portion of participants with Post-Acute Sequelae of SARS-CoV-2 Infection (PASC)
      - a. PASC assessment monthly after enrollment for 6 months to 12 months with the "Questionnaire to characterize long COVID." (Appendix G).[62]"

**Statistical Analysis Plan**

- No changes to the Statistical Analysis Plan have been made since unblinding.
- The SAP was emailed to the DSMB on Feb 14, 2022, before unblinding to the primary outcome on Feb 15, 2022.
- The outcome assessors, patients, care providers and all investigators except the unblinded statistician and graduate student assistant still remain blinded to individual treatment allocation
- PASC is listed as an efficacy outcome in the SAP in section 5.1
- Section 6.4 gives details about how PASC will be analyzed

**Overview changes regarding Long Covid or PASC on Clinical Trials.gov:**

**1.** On clinicaltrials.gov on May 3, 2021, this had been added to the study description:
"5. To understand if any of the active treatment arms prevent long-covid syndrome, PASC (post-acute sequelae of SARS-CoV-2 infection)."

**2.** On clinicaltrials.gov on May 17, 2021 it had been added under primary outcome measures:
"Post-Acute Sequelae of SARS-CoV-2 Infection (PASC) Questionnaire
[ Time Frame: 6 and 12 months ]
PASC assessment will be conducted monthly after enrollment for 6 months to 12 months with the Questionnaire to characterize long COVID. Outcome is reported as the percent of participants who report PASC any symptoms."

**3.** On clinical trials.gov on Sept 30, 2021, it had been moved down to secondary outcome measures:

"Post-Acute Sequelae of SARS-CoV-2 Infection (PASC) Questionnaire
[ Time Frame: 6 and 12 months ]
PASC assessment will be conducted monthly after enrollment for 6 months to 12 months with the Questionnaire to characterize long COVID. Outcome is reported as the percent of participants who report PASC any symptoms."

4. On Clinical trials.gov on Jan 20, 2022 (before enrollment finished), it was still in the study description and still a secondary outcome. The protocol was also uploaded to clinicaltrials.gov in Jan 2022 before enrollment was complete:

"Portion of participants with Post-Acute Sequelae of SARS-CoV-2 infection (PASC)
[ Time Frame: 6 and 12 months ]
PASC assessment will be conducted monthly after enrollment for approximately 9 months with the Questionnaire to characterize long COVID."

**Supplemental Table 5.**

**COVID-OUT Study Team**

| Name | Institute | Location |
|---|---|---|
| Blake Anderson | Emory | Atlanta, GA |
| Riannon C Atwater | University of Colorado | Aurora, CO |
| Nandini Avula | University of Minnesota | Minneapolis, MN |
| Kenny B Beckman | University of Minnesota | Minneapolis, MN |
| Hrishikesh K Belani | Olive View - UCLA | Sylmar, CA |
| David R Boulware | University of Minnesota | Minneapolis, MN |
| Carolyn T Bramante | University of Minnesota | Minneapolis, MN |
| Jannis Brea | Northwestern University | Chicago, IL |
| Courtney A Broedlow | University of Minnesota | Minneapolis, MN |
| John B Buse | University of North Carolina | Chapel Hill, NC |
| Paula Campora | University of Minnesota | Minneapolis, MN |
| Anup Challa | Vanderbilt University | Nashville, TN |
| Jill Charles | University of Minnesota | Minneapolis, MN |
| Grace Christensen | University of Minnesota | Minneapolis, MN |
| Theresa Christiansen | M Health Fairview | Minneapolis, MN |
| Ken Cohen | Optum | Minnetonka, MN |
| Bo Connelly | University of Minnesota | Minneapolis, MN |
| Srijani Datta | University of Minnesota | Minneapolis, MN |
| Nikita Deng | University of Colorado | Aurora, CO |
| Alex T Dunn | Hennepin Healthcare | Minneapolis, MN |
| Spencer M Erickson | University of Minnesota | Minneapolis, MN |
| Faith M Fairbairn | University of Minnesota | Minneapolis, MN |
| Sarah L Fenno | University of Minnesota | Minneapolis, MN |
| Daniel J Fraser | University of Minnesota | Minneapolis, MN |

| | | |
|---|---|---|
| Regina D Fricton | Feinberg School of Medicine, Northwestern | Chicago, IL |
| Gwen Griffiths | University of Minnesota | Minneapolis, MN |
| Aubrey A Hagen | University of Minnesota | Minneapolis, MN |
| Katrina M Hartman | University of Minnesota | Minneapolis, MN |
| Audrey F Hendrickson | Hennepin Healthcare | Minneapolis, MN |
| Jared D Huling | University of Minnesota | Minneapolis, MN |
| Nicholas E Ingraham | University of Minnesota | Minneapolis, MN |
| Arthur C Jeng | Olive View - UCLA | Sylmar, CA |
| Darrell M Johnson | University of Minnesota | Minneapolis, MN |
| Amy B Karger | University of Minnesota | Minneapolis, MN |
| Nichole R Klatt | University of Minnesota | Minneapolis, MN |
| Erik A Kuehl | M Health Fairview | Minneapolis, MN |
| Derek D LaBar | M Health Fairview | Minneapolis, MN |
| Samuel Lee | Feinberg School of Medicine, Northwestern | Chicago, IL |
| David M Liebovitz | Feinberg School of Medicine, Northwestern | Chicago, IL |
| Sarah Lindberg | University of Minnesota | Minneapolis, MN |
| Darlette G Luke | M Health Fairview | Minneapolis, MN |
| Rosario Machicado | Olive View - UCLA | Sylmar, CA |
| Zeinab Mohamud | University of Minnesota | Minneapolis, MN |
| Thomas A Murray | University of Minnesota | Minneapolis, MN |
| Rumbidzai Ngonyama | University of Minnesota | Minneapolis, MN |
| Jacinda M Nicklas | University of Colorado | Aurora, CO |
| David J Odde | University of Minnesota | Minneapolis, MN |
| Elliott Parrens | M Health Fairview | Minneapolis, MN |
| Daniela Parra | University of Minnesota | Minneapolis, MN |
| Barkha Patel | University of Minnesota | Minneapolis, MN |
| Jennifer L Proper | University of Minnesota | Minneapolis, MN |
| Matthew F Pullen | University of Minnesota | Minneapolis, MN |
| Michael A Puskarich | Hennepin Healthcare | Minneapolis, MN |
| Via Rao | University of Minnesota | Minneapolis, MN |
| Neha V Reddy | University of Minnesota | Minneapolis, MN |
| Naveen Reddy | Northwestern University | Chicago, IL |
| Katelyn J Rypka | University of Minnesota | Minneapolis, MN |
| Hanna G Saveraid | University of Minnesota | Minneapolis, MN |
| Paula Seloadji | Olive View - UCLA | Sylmar, CA |
| Arman Shahriar | University of Minnesota | Minneapolis, MN |
| Nancy Sherwood | University of Minnesota | Minneapolis, MN |
| Jamie L Siegart | University of Colorado | Aurora, CO |
| Lianne K Siegel | University of Minnesota | Minneapolis, MN |

| | | |
|---|---|---|
| Lucas Simmons | University of Minnesota | Minneapolis, MN |
| Isabella Sinelli | University of Colorado | Aurora, CO |
| Palak Singh | University of Minnesota | Minneapolis, MN |
| Andrew Snyder | M Health Fairview | Minneapolis, MN |
| Maxwell T Stauffer | St. Olaf College | Northfield, MN |
| Jennifer Thompson | Vanderbilt University | Nashville, TN |
| Christopher J Tignanelli | University of Minnesota | Minneapolis, MN |
| Tannon L Tople | University of Minnesota | Minneapolis, MN |
| Walker J Tordsen | Hennepin Healthcare | Minneapolis, MN |
| Ray HB Watson | University of Minnesota | Minneapolis, MN |
| Beiqing Wu | University of Minnesota | Minneapolis, MN |
| Adnin Zaman | University of Colorado | Aurora, CO |
| Madeline R Zolik | M Health Fairview | Minneapolis, MN |
| Lena Zinkl | M Health Fairview | Minneapolis, MN |
